# Rare Germline Variants in Immune and Drug Target Genes Among Cancer Exceptional Responders

**DOI:** 10.64898/2026.05.14.26352838

**Authors:** Siyuan Chen, Amelia L. M. Tan, Maria C. Saad Menezes, Cassandra L. Perry, Margaret E. Vella, Vinayak V. Viswanadham, Shilpa Kobren, Susanne Churchill, Isaac S. Kohane

## Abstract

**Background:** Cancer treatment response is highly variable, even among patients with the same tumor type and treatment. Exceptional responders (ERs), who are individuals who experience unusually favorable outcomes, provide critical insights into the biological factors driving treatment success. While prior studies have highlighted the role of somatic changes, the contribution of germline rare variants remains underexplored. This study aimed to uncover the genetic underpinnings of exceptional responses by identifying rare, non-silent and predicted deleterious germline mutations enriched among ERs compared to typical cancer patients.

**Methods:** The Network of Enigmatic Exceptional Responders (NEER) project collected clinical and germline whole-genome sequencing (WGS) data from 53 ERs. After quality control procedures and ancestry background checks, 51 ERs were left for final analysis. While non-silent mutations were identified based on allele frequencies and mutation types, multiple pathogenicity predictors were applied for predicted deleterious variants. These were compared to a harmonized and comparable subset from the Pan-Cancer Analysis of Whole Genomes (PCAWG) cohort (n=414) using Fisher’s exact tests. Kaplan-Meier survival analysis applied to evaluate prognostic associations in PCAWG patients. Additionally, Fisher’s exact tests were conducted stratified by cancer type and treatment regimen to identify potential associations between rare germline variants and therapeutic responses.

**Results:** Variants in immune-related genes such as CCL26 and GPRC5D were prevalent, suggesting enhanced immune regulation among ERs. Fourteen genes with non-silent and eight with predicted deleterious mutations showed significantly different frequencies between NEER and PCAWG cohorts (FDR < 0.05). IRX3 emerged as a protective gene enriched in ERs, whereas OR6B2 was associated with poor survival in PCAWG lung cancer patients. Moreover, rare non-silent germline variants in drug target genes were enriched among ERs treated with cisplatin and doxorubicin, implicating altered DNA repair and drug-binding mechanisms in their remarkable outcomes.

**Conclusions:** This study reveals a distinctive germline mutation landscape in exceptional cancer responders, marked by immune-related and drug-target-associated variants that may enhance therapy response and prolong survival. The findings highlight potential novel prognostic biomarkers, such as IRX3 and OR6B2, providing a foundation for developing personalized cancer treatments informed by rare genetic variation.

## Introduction

Cancer treatment response is assumed to depend on molecular features of cancer, including both germline and somatic variants, epigenetic alterations, tumor microenvironment, etc. Even though a large number of biomarkers and companion diagnostics methods have been discovered, the impact on outcomes has been incremental^1–5^. We still lack knowledge on why some cancer patients show far better responses than others. Cancer exceptional responders (ERs) are prismatic examples of unexplained variation in response. They have unusual positive responses to cancer therapies that are not effective for most other patients. They are so rare that they are not likely to be identified by most drug trials^6^. Previously, these patients were considered as idiosyncratic outliers and understudied. Recent studies suggest that ERs may share common characteristics and reveal the somatic genetic basis of ERs in several cases^7,8^. These somatic variants may contribute to exceptional responses to treatments with hypothesized mechanisms including DNA damage, signaling, and the immune response^9^. The contribution of germline variants has received comparatively less attention and is mostly found in case reports. For example, an entire BRCA1 germline gene deletion was found in a patient with recurrent ovarian cancer, who showed an exceptional response to Olaparib^10^.

Two patients with breast cancer and different germline PTEN mutations both showed an exceptional response to the AKT inhibitor, capivasertib^11^. The Network of Enigmatic Exceptional Responders (NEER) project created an ER registry open to all such patients across all trials^12^. The project collected clinical information, germline whole genome sequencing (WGS) data from 53 ERs to determine what genetic germline variations could contribute to exceptional responses. In a previous study, we focused on the effects of germline common variants and reported that ERs exhibit distinct genetic risks for autoimmune disorders, suggesting that enhanced inherited immune responses may contribute to their exceptional outcomes ^13^. However, here, we investigated the hypothesis that rare variants, specifically those that are in protein-coding regions of the gene, contribute to exceptional responses. We compared the frequency of ERs carrying at least one non-silent or predicted deleterious mutation for each gene to that of cancer patients in PCAWG, a pan-cancer WGS dataset^14^. We found 14 genes with non-silent mutations and 8 genes with predicted deleterious mutations that were significantly overrepresented in NEER. To further investigate the functional and clinical implications of these findings, we conducted survival analyses within the PCAWG cohort to determine whether these mutations could predict patient prognosis and explored their distribution across specific tumor types. Additionally, we examined the presence of rare non-silent mutations in drug target genes among NEER ERs to uncover potential mechanisms of therapy response. These analyses provide insights into the genetic underpinnings of exceptional responses and highlight candidate genes that may serve as prognostic biomarkers or therapeutic targets.

## Methods

### Data acquisition

NEER participants were selected from a group of interested US-based applicants aged 18 and above based on a demonstration of an exceptional response to cancer therapies. This was generally considered to be 2 standard deviations outside of the most recently available survival rates. Individuals were also selected in cases of unique or minimal treatment, or outdated treatment with poorer chances of survival. Each new applicant was reviewed by the study team, and in borderline cases by a member of disease-specific oncology experts was consulted to determine an exceptional response. Participants provided a whole blood sample for DNA analysis, sequenced by the Broad Institute for WGS with 30X mean coverage (Supplementary Table 1). WGS data was processed through the CGAP portal developed by Harvard’s Department of Biomedical Informatics and Brigham Genomic Medicine. The Institutional Review Board (IRB) of Harvard Faculty of Medicine gave ethical approval for this work. Individuals consented to participation using a written consent form that was signed digitally. All relevant ethical regulations including the Declaration of Helsinki have been compiled. All participants provided informed consent for themselves – none were deceased at the time of enrollment. The study was publicly posted on the people-powered medicine website (https://peoplepoweredmedicine.org/neer).

### Variants filtering and predicted deleterious variants prediction

At the variant level, VQSR and GT>20 were applied for quality control in both datasets. Variants on the sex chromosomes were excluded. The NEER dataset was from the GRCh38 genome assembly, and the PCAWG dataset was from GRCh37. The Liftover algorithm was used to convert the dataset from one genome assembly to another to make them more comparable^6^. A Hardy-Weinberg filter (p > 0.001) was applied because of the difference in the heterozygous/homozygous ratio between the two cohorts (Sup Fig.1).

**Figure 1.**
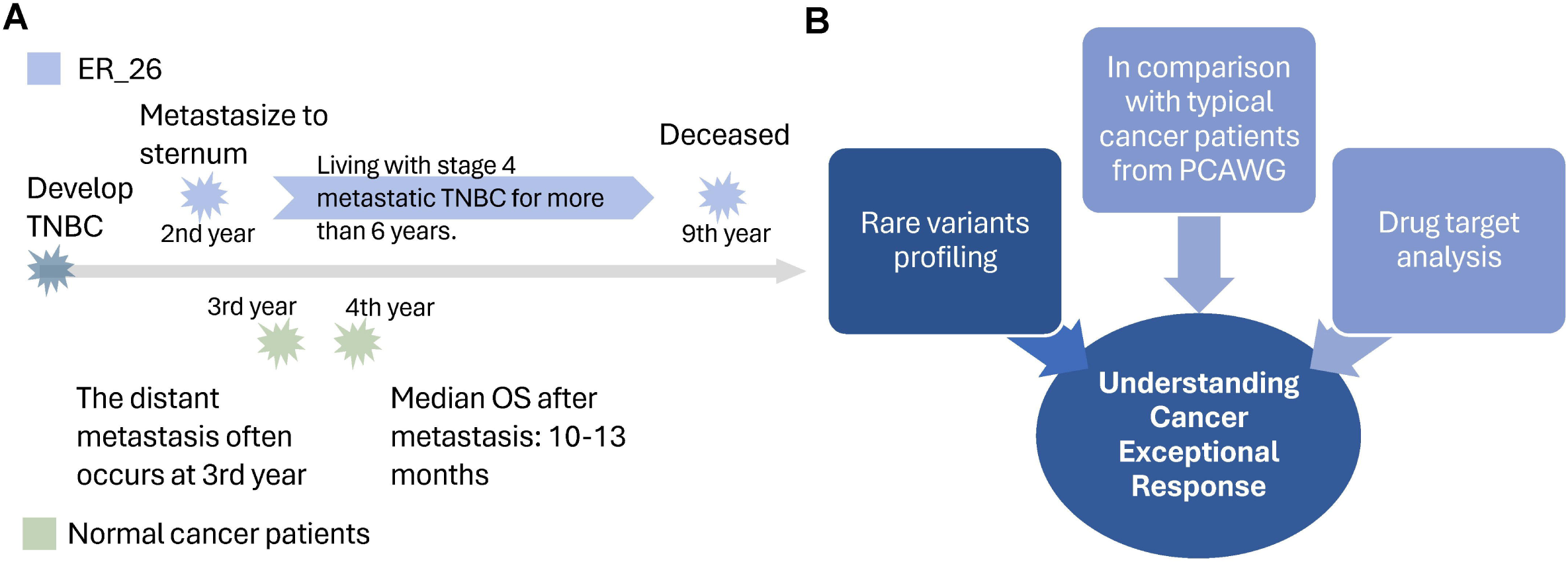
Study design for understanding cancer ERs. A) An example of the difference between typical TNBC patients and one TNBC ER. The blue part showed the life of ER_26 after she was diagnosed with TNBC while the green part showed some epidemiological statistics of TNBC. B) A diagram showed the overall study design to reveal differences between cancer ERs and typical cancer patients.

All variants were annotated through ANNOVAR^15^. Variant filtering was done based on ACMG criteria and Fan S, et al. ^16,17^. Only variants located in exon regions and within ±2 bp sites of exons, the latter potential splicing sites, were kept. Because ERs are rare and the comparison was based on the European population, variants with allele frequency ≥ 0.01 in gnomAD were removed. All synonymous mutations were then removed and the remaining variants were considered as a set of non-silent variants (Sup Fig.1).

A pathogenicity prediction was conducted among all non-silent variants. All variants located in splicing sites, stopgain, stoploss variants, and indels were identified as predicted deleterious variants. To predict the pathogenicity of missense variants, 5 predictive tools were applied, including SIFT^18^, PolyPhen-2^19^, MutationTaster^20^, CADD^21^ and DANN^22^. Missense variants being damaging/unknown in 3 or more softwares were identified as predicted deleterious variants (Sup Fig.1).

### Comparison between NEER and PCAWG

To obtain a comparable control set, an ancestry check was performed through PCA based on all shared variants between NEER ERs and PCAWG typical cancer patients. Two ERs that are not of European ancestry were considered as outliers and removed^13^. As the remaining 51 ERs from NEER were situated within a European population, only the European population patients in the PCAWG cohort were retained for analysis. Patients from the PCAWG cohort were matched to NEER ERs based on tumor types and demographic data^13^. Finally, the sample size of harmonized typical cancer patients from the PCAWG dataset was 414.

A gene-level comparison was conducted only with variants called in both cohorts. Fisher’s exact tests for each gene on the frequency of patients carrying at least one non-silent or predicted deleterious mutation were performed. Another Fisher’s exact test on rare synonymous mutations was done as a negative control to remove systematic bias from the comparison. False discovery rate (FDR) was applied for multiple testing corrections. Only genes with FDR < 0.05 in the non-silent or predicted deleterious mutation comparison and with FDR > 0.10 in the negative control comparison were considered to be significantly different between the two cohorts.

### Survival analysis

A Kaplan-Meier (KM) analysis was performed among all PCAWG European patients. Patients with missing data in vital status were excluded. For the non-silent mutation comparison, patients were divided into two groups based on whether they carried at least one non-silent mutation in the significant genes with an odds ratio > 1. Similarly, patients were divided into two groups based on whether they carried at least one predicted deleterious mutation in the significant genes with an odds ratio > 1 for the predicted deleterious mutation comparison.

### Statistical analysis

Fisher’s exact test was applied to the comparison between NEER and PCAWG, as well as other 2×2 tables in the study. Log-rank test was used to compare the difference in survival time in the KM curve. All tests were two-tailed tests at an α < 0.05 level. The Benjamini-Hochberg Procedure was applied when multiple testing corrections were required, and FDR was calculated.

## Data Accessibility

All SNP data and diagnoses produced in the present study are available upon request to the authors. The PCAWG data can be obtained through the ICGC Data Portal (https://dcc.icgc.org/pcawg).

## Results

### Study design

We recruited a cohort of 53 cancer ERs with different cancer types, from whom germline whole genome sequencing data were acquired. As described in the previous paper, two ERs were considered as outliers due to non-European ancestry and thus were removed from the analysis ^13^. The NEER ERs are so rare by virtue of their extremely long survival time. For example, triple-negative breast cancer (TNBC), accounting for about 15% breast cancer^23^, is highly invasive and is not sensitive to endocrine therapy or molecular targeted therapy. Distant metastasis often occurs in the 3rd year after diagnosis with TNBC^24^. The 5-year mortality rate is 40% within the first 5 years after diagnosis, but the median overall survival of metastatic TNBC is only 10-13 months^25,26^ (Fig.1 A). In contrast, ER_26 in NEER was diagnosed with TNBC in 2011 and had metastases on her sternum at the end of 2013. However, she lived with metastatic Stage 4 TNBC basically as a chronic illness until her death in 2020, which is far longer than the median 10-13 months survival time in metastatic TNBC patients (Fig.1 A). The detailed clinical data for each ER could be found in Supplementary Table 1.

As NEER ERs are definitionally rare among cancer patients, we were curious about the factors accounting for the exceptional responses. We hypothesized that a single rare non-silent mutation on a specific gene could contribute to an exceptional response. To reveal the difference between ERs and typical cancer patients, we not only profiled all rare non-silent mutations and predicted deleterious mutations in NEER ERs, but also compared the ERs to typical cancer patients in PCAWG ^14^, a pan-cancer whole genome sequencing dataset. We also investigated whether there were rare non-silent mutations on known drug targets that could shed light on the relationship between gene mutations and drug responses more directly (Fig.1 B).

### Variant summary for NEER ERs

We generated a variant summary for all rare predicted deleterious variants in NEER. The analysis of variant distributions across samples revealed a median of 233 variants per patient (Fig.2 A). Most predicted deleterious variants are missense mutations, followed by in-frame deletions and nonsense mutations, with significant variability across samples (Fig.2 B, Sup Fig. A). More deletions were found compared to the insertions (Sup Fig.2 B). Among all predicted deleterious point mutations, transitions between C and T occur most while transversion from T to A does not often happen (Fig.2 C).

**Figure 2.**
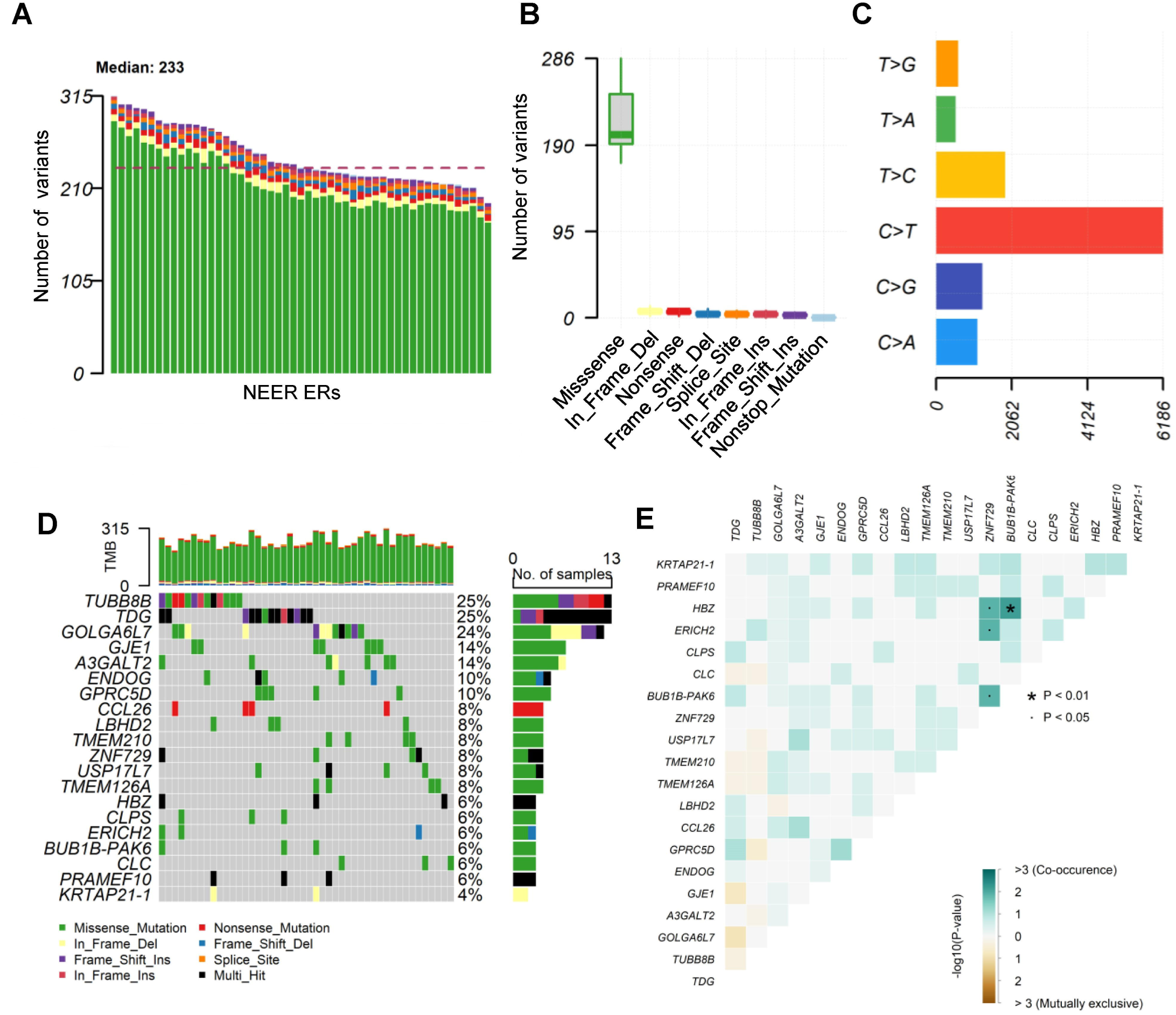
Germline predicted deleterious variant summary for NEER. A) Number of germline predicted deleterious variants identified in each cancer ER. ERs are sorted by number of variants. B) Box plot summarizing the distribution of variant classifications across samples. The plot indicates the median, quartiles, and outliers, emphasizing the prevalence of different mutation types. C) Bar chart illustrating the frequency of single nucleotide variant (SNV) classes based on base substitutions. D) Matrix plot showing mutation types across top mutated genes normalized by gene length. Different color indicates different mutation types, each column is one ER and each row is one gene. The color legend was shared for panel A), B) and D). Number of ERs carrying mutations for a given gene is visualized by the bar chart on the right. E) Heatmap depicting the co-occurrence and mutual exclusivity of mutations between gene pairs of the top mutated genes in panel D. Those genes likely to be mutated simultaneously are labeled in green while those genes less likely to be co-mutated are labeled in brown. Significant interactions are marked. ‘.’ indicates P < 0.05 and ‘*’ indicates P<0.01.

The mutation landscape of the top 20 mutated genes was listed (Sup Fig.2 C). However, gene length can affect gene mutation frequencies. Therefore, we further adjusted the mutation frequency of genes by gene length, with the resulting top-ranked 20 genes after gene-length correction shown (Fig.2 D). *TUBB8B* is the most mutated gene, which disrupts microtubule dynamics and can affect cell division, potentially contributing to cancer progression. 8 of 20 genes (A3GALT2, ENDOG, GPRC5D, CCL26, ZNF729, HBZ, CLC, PRAMEF10) were reported to have immune-related or cellular signaling function, which suggests some potential differences in immune background or immune regulation between ERs and typical cancer patients, which is consistent with our findings in the previous study ^13^. Around 25% of ERs carried a stop-gain mutation on CCL26. We then examined mutation co-occurrence among these genes. Among the top mutated genes without gene-length adjustment, HSPG2 and HMCN2 are mutated simultaneously in ERs (Sup Fig.2 D). HBZ and BUB1B-PAK6 are most likely to be concurrently mutated among the top mutated genes after gene-length adjustment (Fig.2 E).

### Genes with different mutation frequencies between NEER and PCAWG

We compared the frequency of patients carrying at least one non-silent or predicted deleterious mutation between NEER and PCAWG. The datasets were harmonized as described in our previous paper ^13^ (see methods). ERs in NEER were older and of later cancer stages compared to typical cancer patients, but they lived about 4 times longer (Table1).

14 non-silent and 8 predicted deleterious mutation significant genes were identified as having significantly different frequencies (Supplementary Table 2, Supplementary Notes). We refer to these genes as significantly differential frequency (SDF) genes. 7 of the SDF genes were shared in both comparisons (Fig.3 A). In the non-silent mutation comparison, 24 of 51 ERs carried at least one non-silent mutation in the significant genes and 3 of them carried two or more mutated genes (Fig.3 B). Patients without mutated genes include SDF genes with an odds ratio (OR) < 1. *IRX3*, the only SDF gene with OR > 1 in both comparisons, was reported to be an oncogene in T cell acute lymphoblastic leukemia and has tumor-promoting potential ^27,28^(Fig. 3 C,D). The annotation of the SDF genes include structural constituents of cytoskeleton, peptide binding, and MHC class II mediated immune activity (Sup Fig.3). For ERs carrying mutations on three genes with OR > 1 from the non-silent mutation comparison, three ERs carried non-silent mutations on two of these genes (Fig.3 E). For those SDF genes with OR < 1, most ERs do not have any non-silent or predicted deleterious mutations on these genes. Three ERs carried non-silent mutations on HRNR and two different ERs carried non-silent mutations on OR6B2 and CDC14C separately. Additionally, two ERs carried predicted deleterious mutations on EPPK1(Sup Fig 4). Detailed functional description of the mutations identified on these three genes was listed in Supplementary Table 3. Three rare non-silent variants identified on AOC1 (rs148613441, rs200206446, rs200630234) are associated with type 1 diabetes in the European population, which was consistent with our previous findings that ERs have higher Type 1 diabetes risks compared to typical cancer patients^13^.

**Fig 3.**
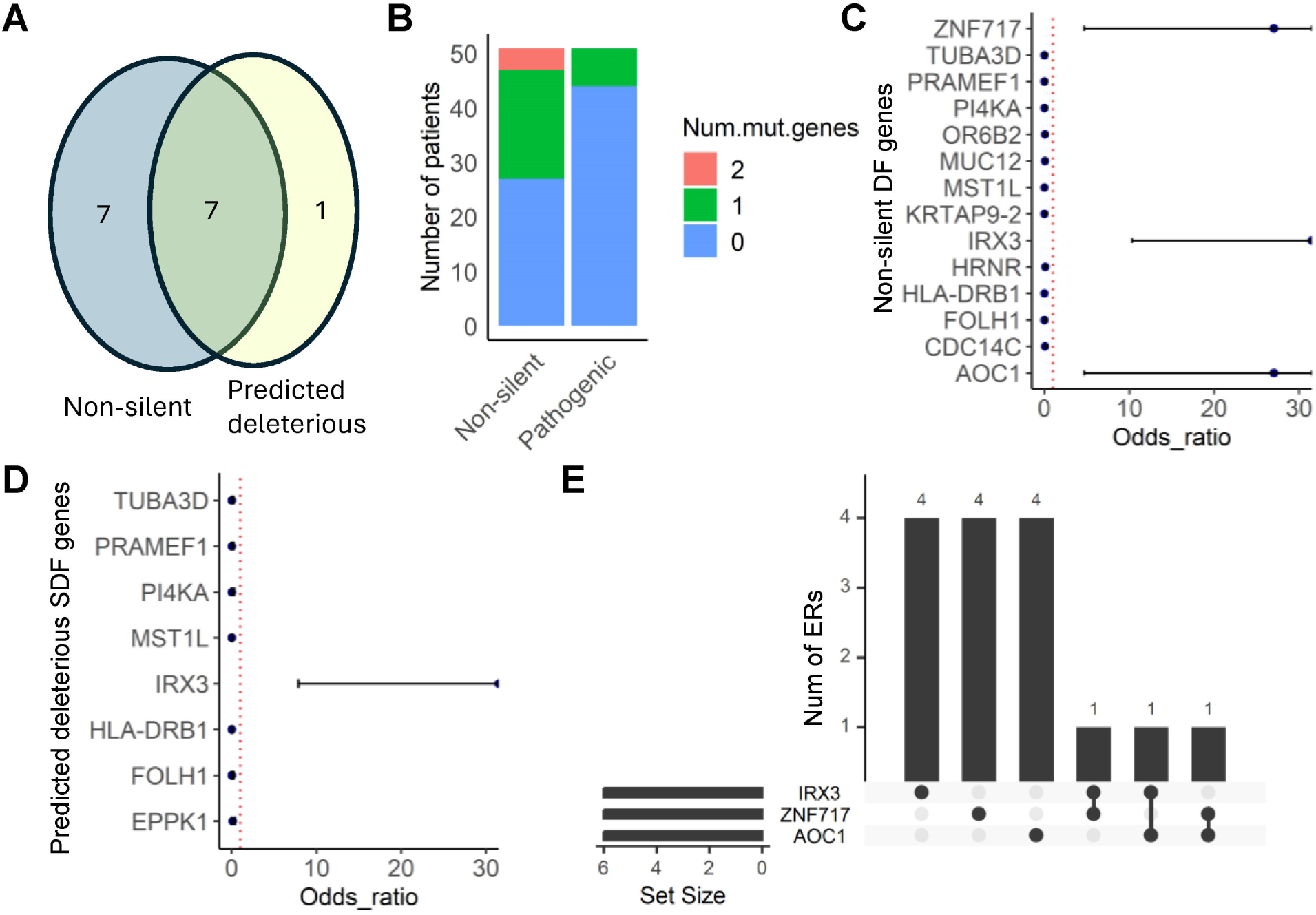
Comparison of gene mutation frequency between NEER and PCAWG. A) Venn diagram comparing the number of significant genes with non-silent mutations and predicted deleterious mutations. B) Bar plot showing the number of patients with non-silent and predicted deleterious mutations on SDF genes respectively. C,D) Dot plots displaying the odds ratios for SDF genes with non-silent mutations and predicted deleterious mutations respectively from our comparison. E) Upset plot visualizing the intersection of patients with non-silent mutations in IRX3, ZNF717 and AOC1. The number of ERs with mutations on these genes was shown in the bar chart.

**Figure 4:**
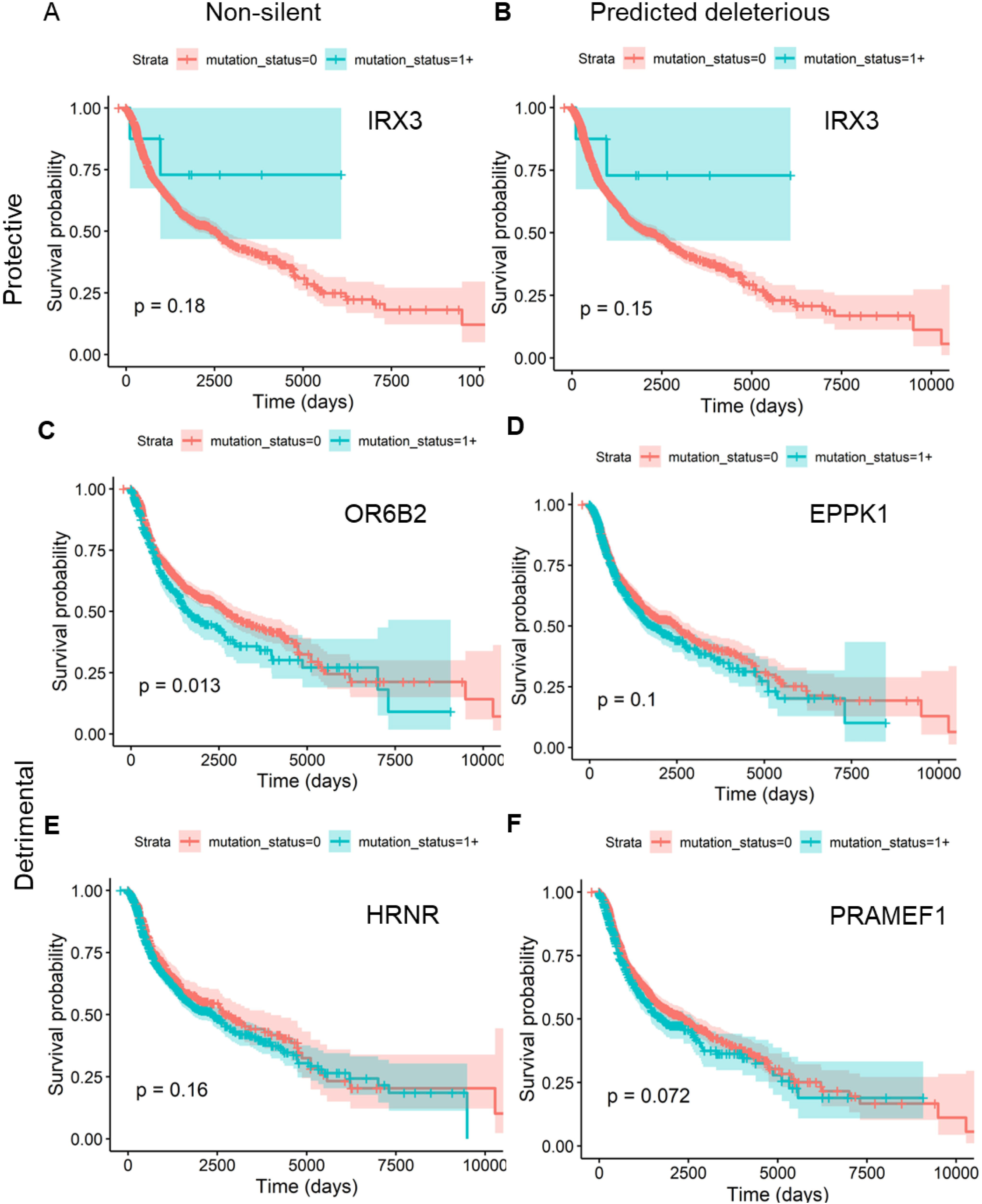
Survival analysis based on SDF genes among PCAWG patients. KM survival curve for 6 significant genes in the comparison. The blue curve refers to patients carrying at least one non-silent or predicted deleterious mutation on the given gene, while the red curve refers to patients without mutations. Protective genes indicate SDF genes with odds ratios > 1 and detrimental genes are SDF genes with odds ratios < 1. A)C)E) IRX3, OR6B2 and HRNR are from the non-silent comparison and only non-silent mutations are considered in the survival analysis. B)D)F) IRX3, EPPK1 and PRAMEF1 are from the predicted deleterious comparison and only predicted deleterious mutations are considered in the survival analysis.

### Survival analysis within all PCAWG European patients

As the primary difference between ERs and typical cancer patients lies in their survival time, we next investigated whether the genes identified earlier could predict pan-cancer prognosis in the full set of PCAWG European patients. To do this, we divided cancer patients into two groups based on whether they carried at least one non-silent mutation or predicted deleterious mutation in a given gene. Genes with an OR > 1 were classified as having protective effects, while genes with an OR < 1 were considered detrimental. Among the PCAWG typical patients, those carrying non-silent mutations in OR6B2 exhibited a significant decrease in survival time, suggesting its role as a detrimental gene (Fig. 4).

We next sought to determine whether the SDF genes identified from our comparison have specific tumor type associations. We analyzed the distribution of non-silent mutations on SDF genes across tumor types among PCAWG patients. Non-silent mutations in OR6B2, PI4KA, and CDC14C were more frequently found in lung cancer patients (Fig.5 A, Sup Fig.5). To further investigate whether these mutations are associated with differences in patient survival within the same tumor type, we performed survival analyses within individual tumor types among PCAWG patients. Consistent with the tumor-progressing roles of OR6B2 observed between ERs and typical cancer patients, lung cancer patients in the PCAWG cohort who harbored non-silent mutations in OR6B2 had significantly shorter survival times compared to those without mutations in this gene (Fig.5 B), while no significantly different in survival time were observed on the other two gene among lung cancer patients (Sup Fig.6). Additionally, among PCAWG patients, mutations in PI4KA were enriched in ovarian cancer patients, while TUBA3D mutations were more common in skin cancer patients (Sup Fig.5). Although the difference was not statistically significant, there was a trend towards shorter survival times in PCAWG skin cancer patients with TUBA3D non-silent mutations compared to those without (Sup Fig.6). These analyses within the PCAWG cohort indicate that non-silent mutations in certain SDF genes are enriched in specific tumor types and, in some cases, are associated with differences in survival outcomes within those tumor types.

**Figure 5.**
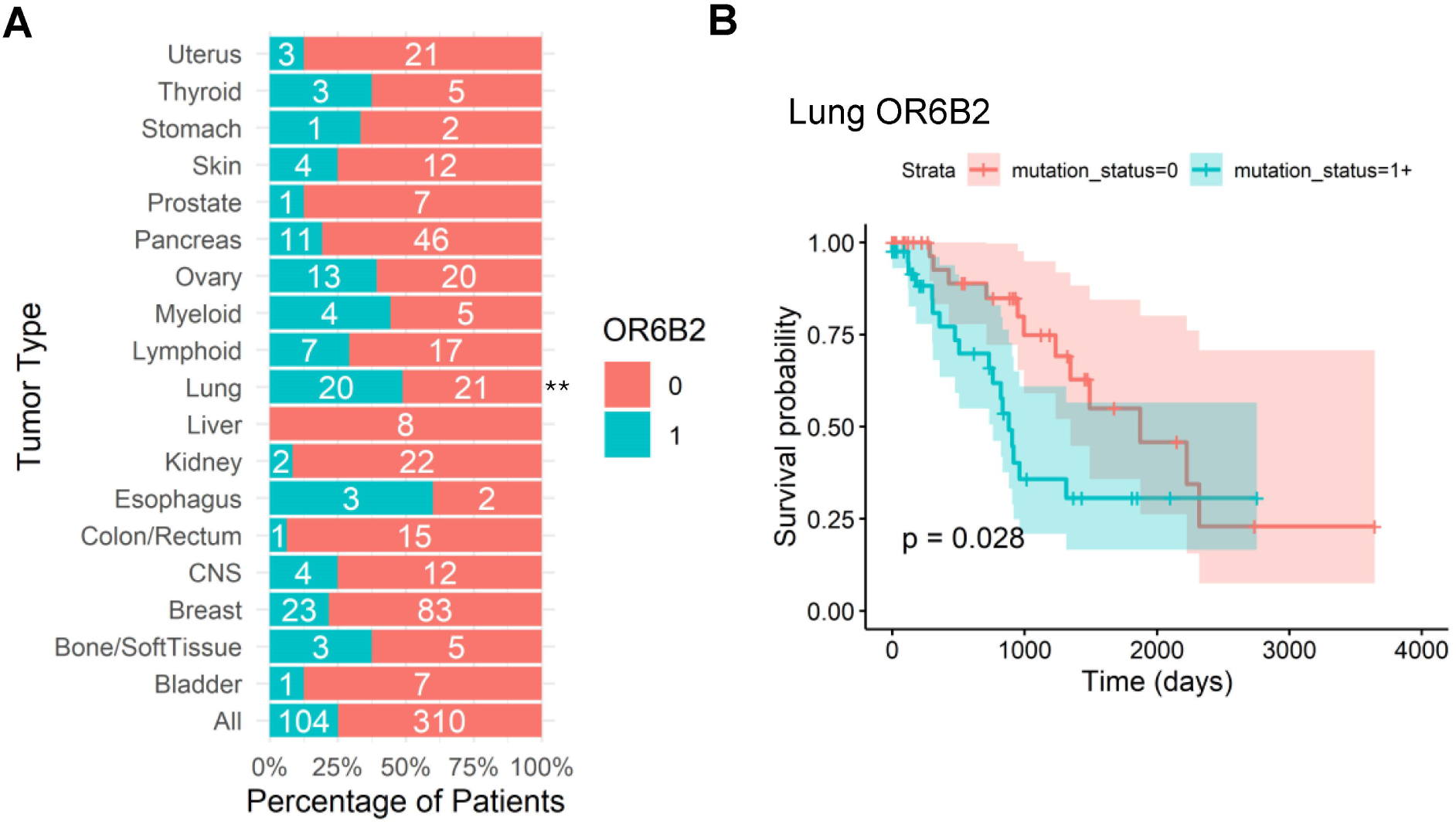
OR6B2 is a potential prognostic marker in lung cancer. A) Percentage of PCAWG patients with various tumor types harboring rare non-silent mutations in OR6B2. “1” indicates the presence, and “0” the absence, of such mutations. An asterisk (*) denotes a significantly different proportion of mutated patients in the indicated tumor type compared to the overall cohort. B) KM survival analysis of PCAWG lung cancer patients stratified by the presence (mutation status = 0) or absence (mutation status = 1+) of rare non-silent OR6B2 mutations.

**Figure 6.**
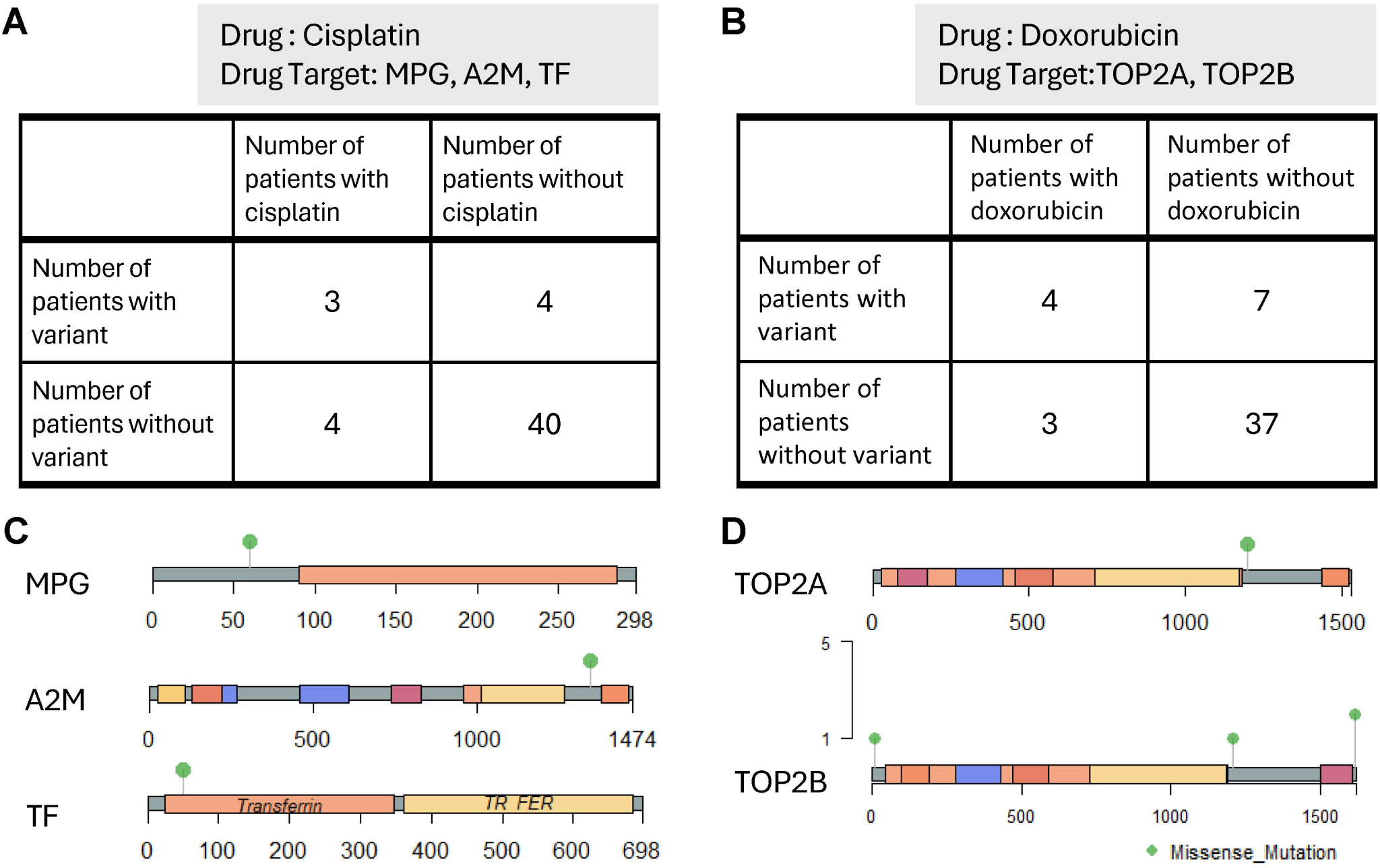
Elaboration from hits in Drug targets analysis. A & B) 2×2 tables showing the number of ERs with mutated or non-mutated drug target genes and the number of ERs prescribed or not prescribed with the drug. C&D) Lolliplot showing where the variants were located on the drug target genes.

### Rare non-silent variants were found on target genes of cisplatin and doxorubicin

As therapy is another important factor contributing to patients’ survival, we next questioned whether there were rare non-silent mutations on the targets of drugs assigned to ERs. NEER ERs went through different therapies, including antiangiogenic therapy, chemotherapy, hormone therapy, immunotherapy, radiation and targeted therapy. Chemotherapies were assigned to 73.21% of the patients, with antimetabolite and anti-mitotic drugs both given to 18 ERs (Table 2). Except for radiation, paclitaxel was the most common treatment, followed by gemcitabine and carboplatin, which were given to 13, 11 and 9 ERs, respectively (Table 3). Detailed drug assignment data for each patient can be found in the Supplementary Table 4, and we focused on commonly used drugs that were administered to more than 6 NEER ERs.

**Table 1.**
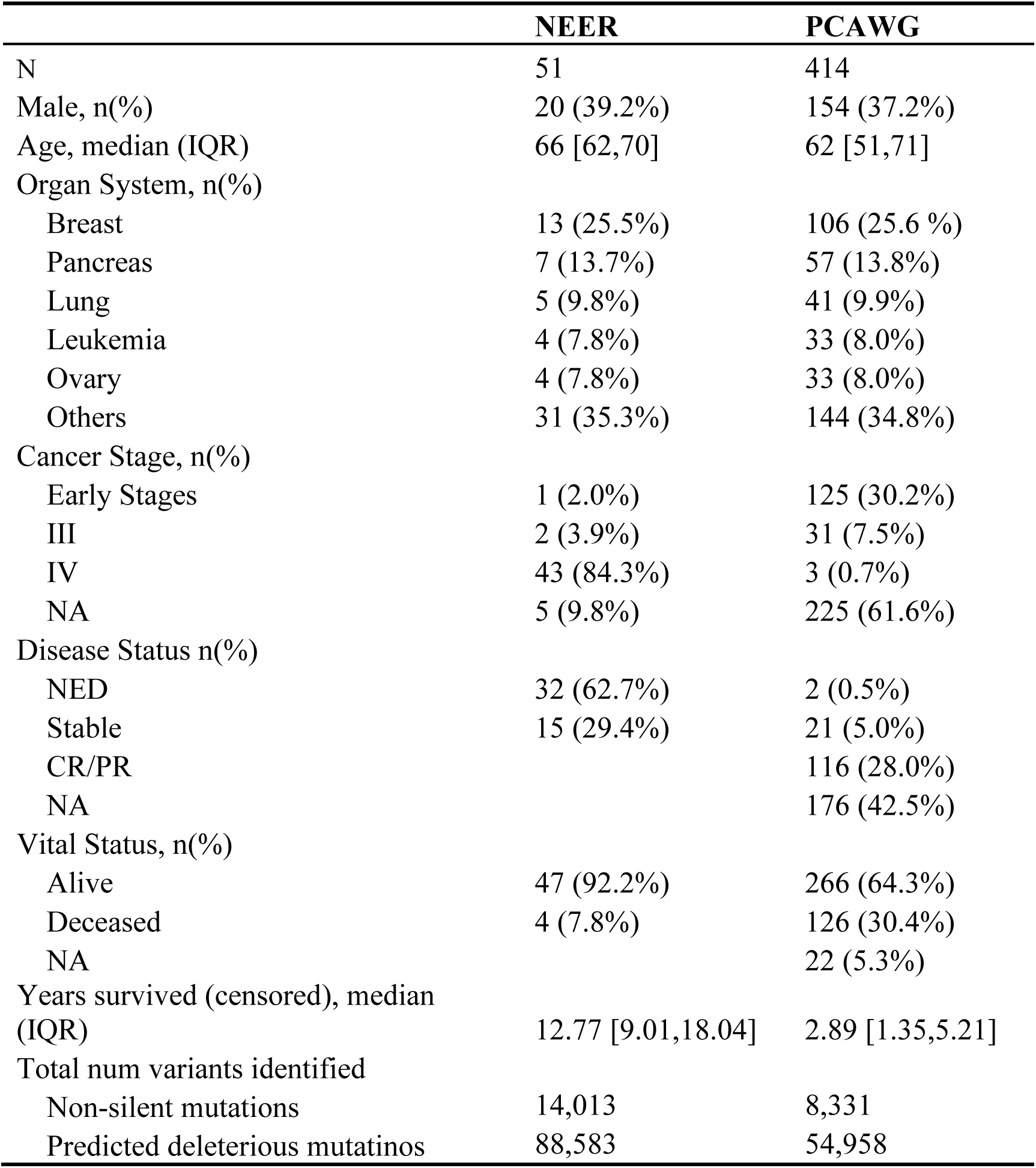
Demographic and clinical summary between NEER and PCAWG patients.

|  | NEER | PCAWG |
| --- | --- | --- |
| N | 51 | 414 |
| Male, n(%) | 20 (39.2%) | 154 (37.2%) |
| Age, median (IQR) | 66 [62,70] | 62 [51,71] |
| Organ System, n(%) |  |  |
| Breast | 13 (25.5%) | 106 (25.6 %) |
| Pancreas | 7 (13.7%) | 57 (13.8%) |
| Lung | 5 (9.8%) | 41 (9.9%) |
| Leukemia | 4 (7.8%) | 33 (8.0%) |
| Ovary | 4 (7.8%) | 33 (8.0%) |
| Others | 31 (35.3%) | 144 (34.8%) |
| Cancer Stage, n(%) |  |  |
| Early Stages | 1 (2.0%) | 125 (30.2%) |
| III | 2 (3.9%) | 31 (7.5%) |
| IV | 43 (84.3%) | 3 (0.7%) |
| NA | 5 (9.8%) | 225 (61.6%) |
| Disease Status n(%) |  |  |
| NED | 32 (62.7%) | 2 (0.5%) |
| Stable | 15 (29.4%) | 21 (5.0%) |
| CR/PR |  | 116 (28.0%) |
| NA |  | 176 (42.5%) |
| Vital Status, n(%) |  |  |
| Alive | 47 (92.2%) | 266 (64.3%) |
| Deceased | 4 (7.8%) | 126 (30.4%) |
| NA |  | 22 (5.3%) |
| Years survived (censored), median (IQR) | 12.77 [9.01,18.04] | 2.89 [1.35,5.21] |
| Total num variants identified |  |  |
| Non-silent mutations | 14,013 | 8,331 |
| Predicted deleterious mutatinos | 88,583 | 54,958 |

**Table 2.**
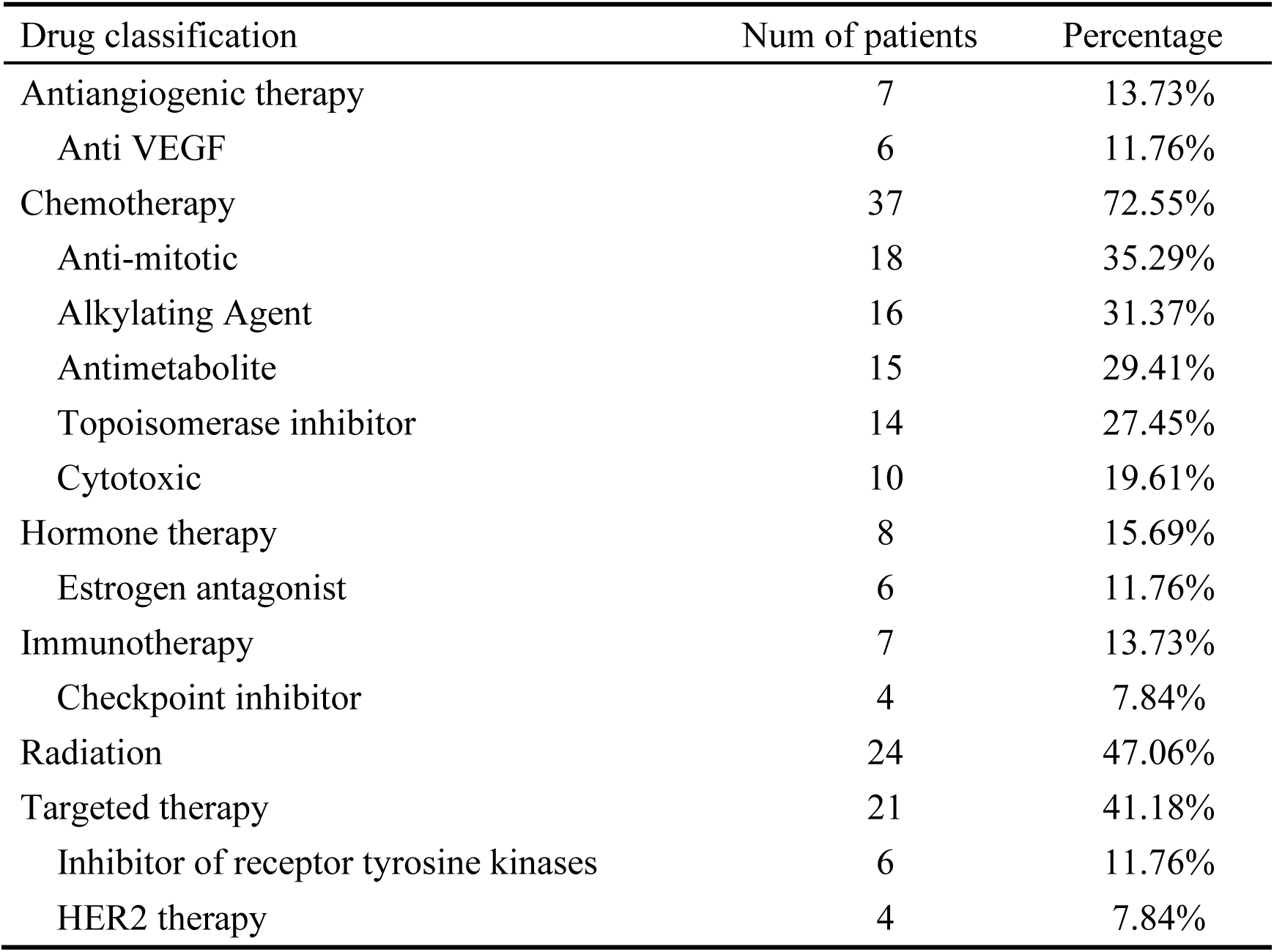
Summary of treatment classes prescribed to NEER ERs.

| Drug classification | Num of patients | Percentage |
| --- | --- | --- |
| Antiangiogenic therapy | 7 | 13.73% |
| Anti VEGF | 6 | 11.76% |
| Chemotherapy | 37 | 72.55% |
| Anti-mitotic | 18 | 35.29% |
| Alkylating Agent | 16 | 31.37% |
| Antimetabolite | 15 | 29.41% |
| Topoisomerase inhibitor | 14 | 27.45% |
| Cytotoxic | 10 | 19.61% |
| Hormone therapy | 8 | 15.69% |
| Estrogen antagonist | 6 | 11.76% |
| Immunotherapy | 7 | 13.73% |
| Checkpoint inhibitor | 4 | 7.84% |
| Radiation | 24 | 47.06% |
| Targeted therapy | 21 | 41.18% |
| Inhibitor of receptor tyrosine kinases | 6 | 11.76% |
| HER2 therapy | 4 | 7.84% |

**Table 3.** Summary of drugs prescribed to NEER ERs.

| Drugs | Total | Level2_classification | Level1_classification |
| --- | --- | --- | --- |
| Radiation (NOS) | 18 | Radiation | Radiation |
| Paclitaxel | 13 | anti-mitotic | Chemotherapy |
| Carboplatin | 9 | Alkylating Agent | Chemotherapy |
| Gemcitabine | 8 | Antimetabolite | Chemotherapy |
| Cisplatin | 7 | Alkylating Agent | Chemotherapy |
| Doxorubicin | 7 | topoisomerase inhibitor | Chemotherapy |
| Fluorouracil | 6 | Antimetabolite | Chemotherapy |
| Irinotecan | 6 | topoisomerase inhibitor | Chemotherapy |
| Bevacizumab | 6 | anti VEGF | antiangiogenic therapy |
| Docetaxel | 6 | anti-mitotic | Chemotherapy |

Cisplatin is a platinum-based chemotherapy agent used to treat various sarcomas, carcinomas, lymphomas, and germ cell tumors, which was assigned to 7 patients in NEER. Moreover, 3 of the patients carried a rare non-silent mutation on the target genes, with OR = 7.0 (P=0.045) among 51 NEER ERs (Fig.6 A). Cisplatin works through three different mechanisms. It can attach alkyl groups to DNA bases, induce DNA damage via the formation of cross-links and cause mispairing of the nucleotides, resulting in the disruption of DNA function and cell death. ER_9 carried a chr16:79563:C:T mutation on *MPG*, which is a repair enzyme that can remove cisplatin adducts (Fig.6 C). It has been reported that cisplatin adducts can titrate *MPG* away from its natural substrates, resulting in higher mutagenesis and/or cell death because of the persistence of *MPG* substrates in DNA and overexpression of *MPG* sensitizes cells to alkylating agent-induced cytotoxicity^29,30^. It is also possible that the rare missense mutation on *MPG* impairs DNA damage repair, leading to neoantigen generation, thereby boosting immune response in the ER. Proteomic studies have shown that *A2M* is one of the cisplatin-binding proteins in human serum ^31,32^. Er-JiaWang, et al. reported that *A2M* was 18.3-fold upregulated on the 5th day in rats after being treated with cisplatin^33^. The rare A2M variant we found (chr12:9072429:C:T) in ER_9 may contribute to the response to cisplatin (Fig.6 C). Another platinum drug target is *TF*, which encodes transferrin, which is reported to bind with cisplatin^34–36^. ER_35 carried a rare missense mutation (chr3:133753659:A:G) on the transferrin domain of *TF* (Fig.6 C), which therefore might affect the binding between transferrin and cisplatin.

Doxorubicin, a cytotoxic anthracycline antibiotic to treat various cancers, was used in 7 NEER patients. Doxorubicin has antimitotic and cytotoxic activity through interaction with DNA in a variety of different ways, including intercalation, DNA strand breakage and inhibition of the enzyme topoisomerase II. 4 of 7 ERs carried at least one rare non-silent mutation on *TOP2A* or *TOP2B*, with an odds ratio of 6.67 (P=0.03) (Fig.6 B). Of the two isoforms of topoisomerase II, the targets of doxorubicin, *TOP2A* appears to be associated with replication and cell division, while *TOP2B* is associated with transcription and chromatin topology^37,38^. Decrease in expression of *TOP2A* affects the extent of DNA damage after treatment with doxorubicin and contributes to poorer efficacy^39^. The rare non-silent TOP2A and TOP2B mutations carried by ERs may account for their better response to doxorubicin.

## Discussion

This study studies the rare genetic variant factors contributing to the outcomes observed in a cohort of long-term cancer survivors. We identified rare non-silent and predicted deleterious mutations, revealed novel mutation patterns, and explored their potential contributions to survival, tumor biology, and therapy response. By comparing the mutation profiles limited to rare non-silent mutations and predicted deleterious mutations to typical cancer patients from PCAWG, we reported germline mutations potentially driving the exceptional response.

Our analysis revealed a distinct mutation landscape in NEER ERs, characterized by a higher prevalence of specific rare non-silent and predicted deleterious mutations. Notably, the predominance of C>T transitions, which have been associated with longer survival in cancer patients ^40^, suggests that the mutation patterns in ERs may be linked to improved outcomes. C>A is associated with a shorter survival time, which is less likely to be observed in ERs ^41^. The identification of immune-related genes among the top mutated genes, including CCL26, GPRC5D, and A3GALT2, underscores the potential role of immune regulation in ERs. Loss-of-function mutations in CCL26, a gene known to promote tumor evasion and metastasis^42^ , were observed in approximately 25% of ERs. This suggests that impaired CCL26 function may disrupt the tumor microenvironment, potentially preventing immune evasion and contributing to long-term survival ^43–45^.

By comparing NEER ERs to PCAWG patients, we identified several genes with significant differences in mutation frequencies. Among these, IRX3, a gene previously implicated as an oncogene in T-cell acute lymphoblastic leukemia ^46^, stood out as the only gene with an OR > 1 in both non-silent and predicted deleterious mutation comparisons. This suggests that IRX3 mutations may paradoxically confer protective effects in certain contexts, potentially through mechanisms such as immune activation or suppression of metastatic potential. Conversely, genes with OR < 1, such as HRNR and OR6B2, may represent targets of selective pressure in typical cancer patients but are rarely mutated in ERs, suggesting a protective role against disease progression. OR6B2, an olfactory receptor was found significantly less frequently mutated in ERs and associated with significantly shorter survival times in lung cancer patients. Previous studies have shown that olfactory receptors may be putative drivers of cancer, but how these genes affect cancer survival is not clear ^47^.

Among NEER ERs, we observed that rare non-silent variants in the relevant drug target genes were particularly enriched among those ERs treated with cisplatin or doxorubicin, suggesting a potential link between these genetic alterations and exceptional responses to treatment. For example, mutations in MPG, a repair enzyme involved in the removal of DNA adducts, may impair DNA damage repair and enhance the cytotoxic effects of cisplatin in ERs. Similarly, mutations in A2M and TF, both of which interact with cisplatin, may influence drug binding and efficacy, thereby contributing to prolonged survival ^31,48^.

Mutations in TOP2A may modulate DNA damage repair and enhance therapeutic efficacy, while mutations in TOP2B could potentially reduce off-target toxicities such as cardiotoxicity ^49,50^. These results suggest that rare germline variants in drug target genes may not only improve treatment efficacy but also mitigate adverse effects, ultimately contributing to the exceptional responses observed in this cohort. Interestingly, most of the rare missense mutations were located in unstructured regions of the genes. Since most rare missense mutations are deleterious or at least function-change^51^, and gain-of-function mutations were enriched in unstructured regions^52^, we proposed that gain of function in drug targets may lead to exceptional responses.

While this study provides insights into the genetic basis of exceptional responses in cancer, several limitations should be considered. First, the sample size of NEER ERs is relatively small, which may limit the generalizability of the findings. The complicated and varied therapeutic context among ERs brings obstacles in interpreting the genetic contributions to drug response. In addition, the functional effects of the identified mutations were inferred based on existing literature and computational predictions, and experimental validation is required to confirm their biological relevance.

Overall, this study highlights the unique genetic landscape of NEER ERs, identifying marker genes where rare non-silent and predicted deleterious mutations may contribute to exceptional survival. The findings underscore the critical role of immune regulation and genetic variants in drug target genes in shaping cancer outcomes. It also provides potential prognostic markers that can improve the clinical survival prediction and drug assignments. These insights provide a foundation for future studies aimed at uncovering the molecular basis of exceptional responses and developing novel therapeutic strategies to improve patient outcomes.

## Data Availability

All data produced in the present study are available upon reasonable request to the authors

## Author Contribution

Author Contributions: Siyuan and Dr. Amelia had full access to all the data in the study and took responsibility for the integrity of the data and the accuracy of the data analysis. They contributed equally to this work as co–first authors.

Concept and design: Siyuan, Amelia, Cassandra, Susanne, Shilpa, Issac.

Acquisition, analysis, or interpretation of data: All authors.

Drafting of the manuscript: Siyuan, Amelia, Maria, Isaac.

Critical revision of the manuscript for important intellectual content: Siyuan, Amelia, Maria, Isaac.

Statistical analysis: Siyuan. Obtained funding: Isaac.

Administrative, technical, or material support: Cassandra, Susanne, Margaret. Supervision: Isaac.

## Acknowledgement

We are grateful to the following expert oncologists who helped us evaluate candidate ERs. Dr, David Fisher, Dr. Alice Shaw, Dr. Mark Johnson, and Dr. Funda Meric-Bernstam. We are also thankful for Professors Shamil Sunyaev and Professors Peter Park’s comments which were helpful in improving the manuscript. Large language models were used to revise the grammatical errors and fluency of the manuscript.

## Competing Interest Statement

The authors have declared no competing interest.

## Funding Statement

This study was funded by 1. The People-Powered Medicine platform was made possible by a grant from the National Institutes of Health Big Data to Knowledge program (Grant #U54H6007963). 2. Moskovitz Fund for Precision Medicine 3. Millenium Pharmaceuticals

## Notes

### Author Declarations

The Institutional Review Board (IRB) of Harvard Faculty of Medicine gave ethical approval for this work.

