## Supplementary Figures and Notes for "Rare Germline Variants in Immune and Drug Target Genes Among Cancer Exceptional Responders"

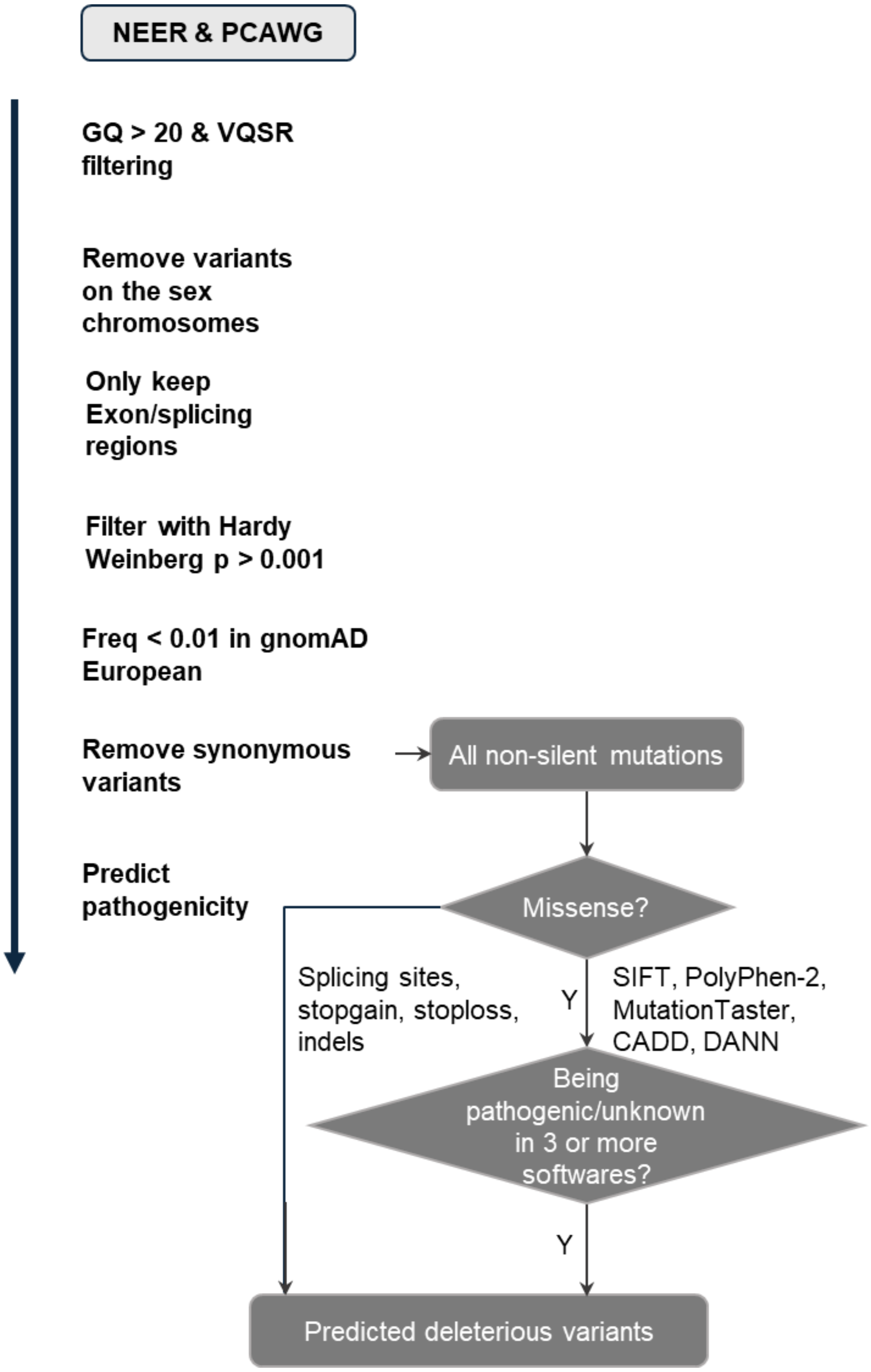


Sup Fig.1 A diagram showing quality controls and filtering steps of the variants.

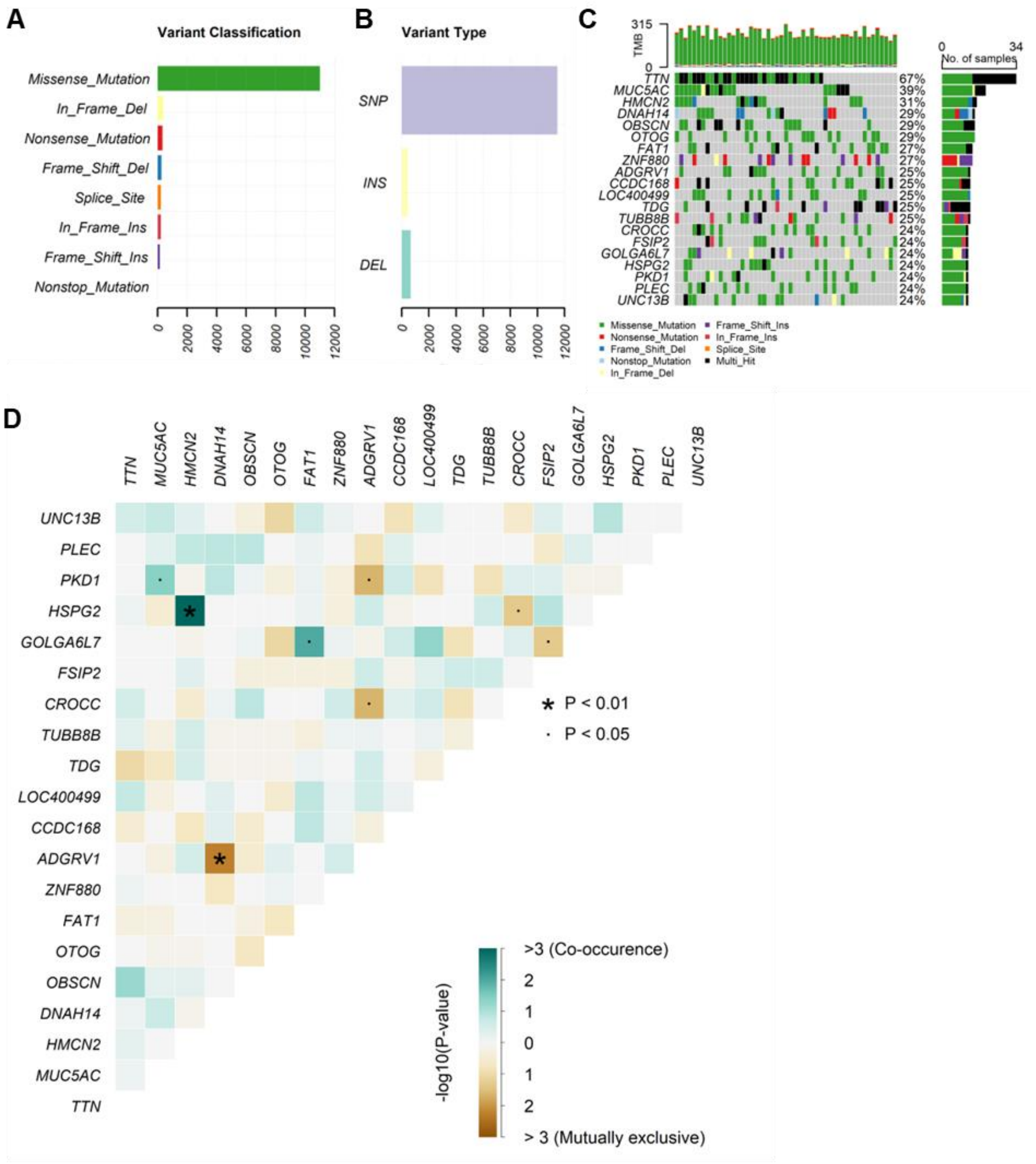


Sup Fig.2 Germline predicted deleterious variants without controlling for gene length

A,B) Number of predicted deleterious variants of different variant classes and types. C) Matrix plot showing mutation types across the top mutated genes with no normalization by gene length. Different colors indicate different mutation types, each column is one ER and each row is one gene. Number of ERs carrying mutations for a given gene is visualized by the bar chart on the right. The color legend was shared for panels A) and C). D) Heatmap depicting the co-occurrence and mutual exclusivity of mutations between gene pairs of the top mutated genes in panel C. Those genes are likely to be mutated simultaneously are labeled in green while those genes that are less likely to be co-mutated are labeled in brown. Significant interactions are marked. '.' indicates P < 0.05 and '*' indicates P<0.01.

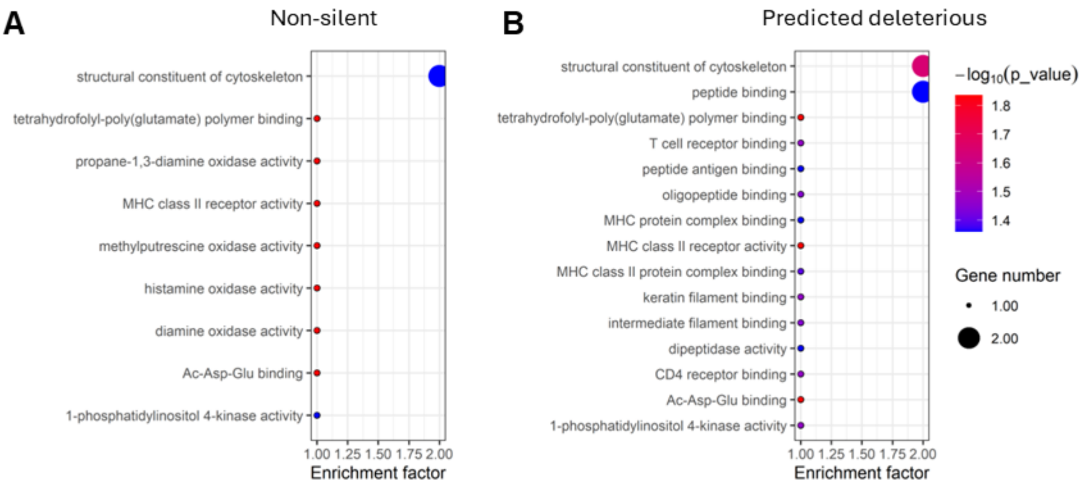


Sup Fig 3 GOBP enrichment for SDF genes from comparison with non-silent and predicted deleterious mutations.

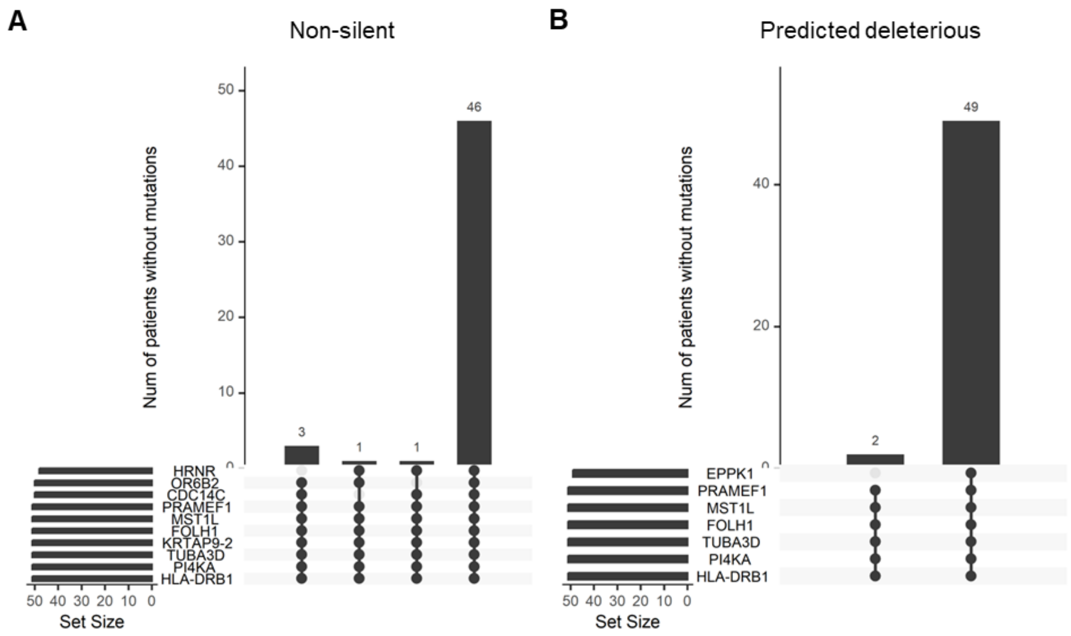


Sup Fig 4 ERs carrying detrimental SDF genes.

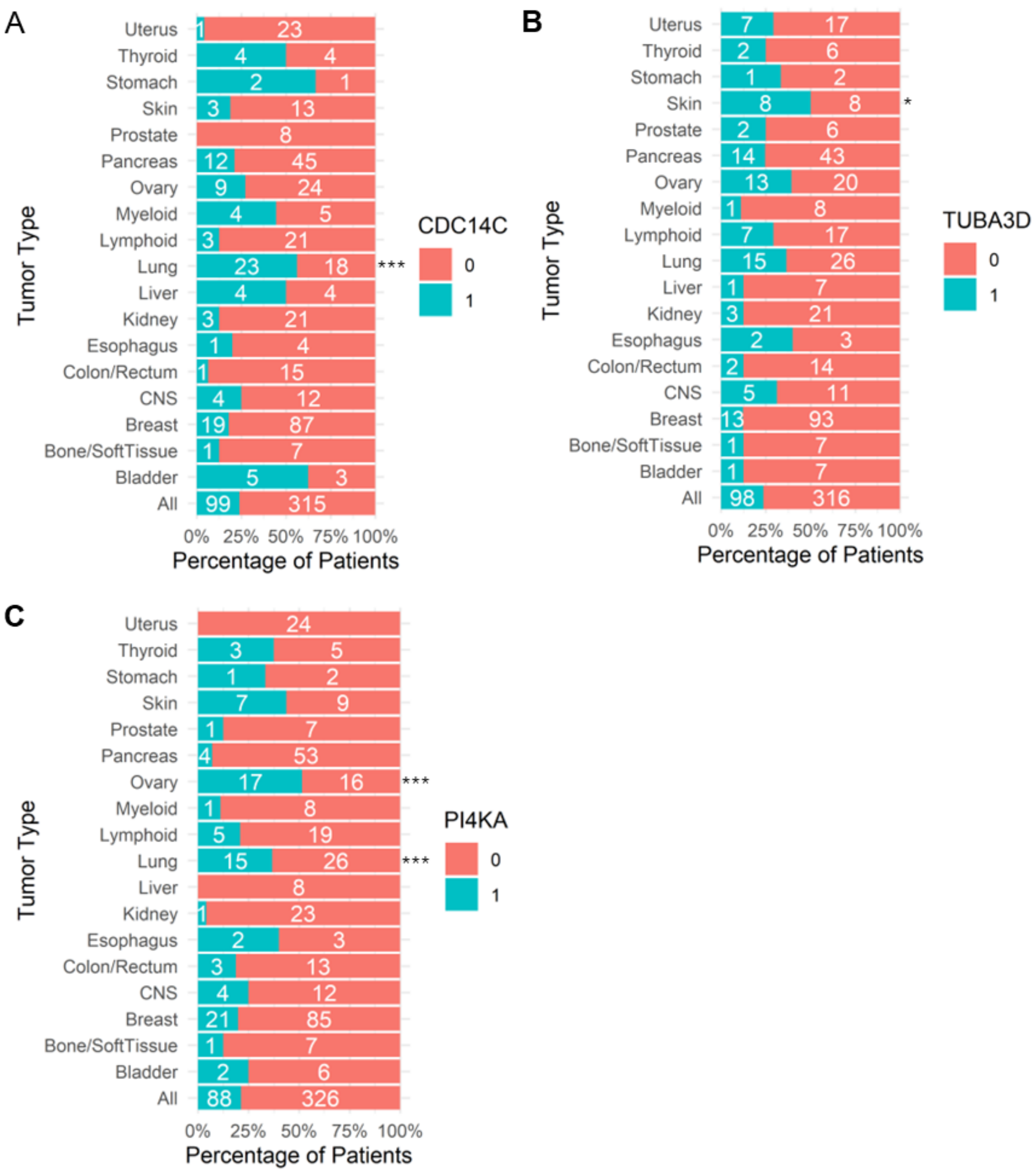


**Sup Fig 5 Proportion of PCAWG patients with rare non-silent mutations in three genes across tumor types**

A) Percentage of PCAWG patients with different tumor types harboring rare non-silent mutations in CDC14C.

B) Percentage of PCAWG patients with different tumor types harboring rare non-silent mutations in TUBA3D.

C) Percentage of PCAWG patients with different tumor types harboring rare non-silent mutations in PI4KA.

For all panels, "1" indicates the presence and "0" the absence of rare non-silent mutations in the respective gene. An asterisk (*) denotes a significantly different proportion of mutated patients in the indicated tumor type compared to the overall cohort.

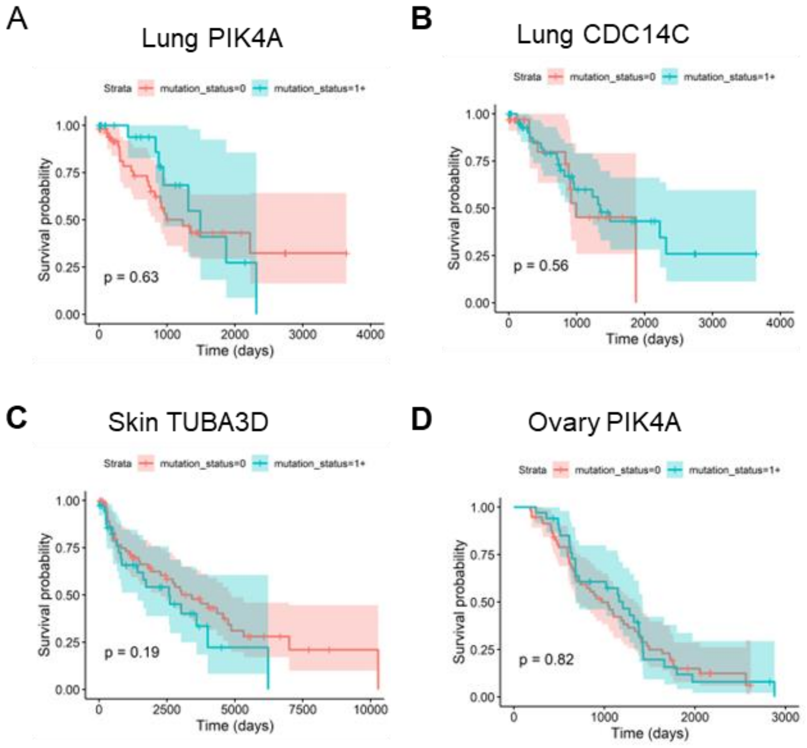


**Sup Fig 6 KM survival analyses of rare non-silent mutations in selected genes across tumor types in PCAWG patients.**

A) KM survival analysis of PCAWG lung cancer patients stratified by the presence or absence of rare non-silent PI4KA mutations.
B) KM survival analysis of PCAWG lung cancer patients stratified by the presence or absence of rare non-silent CDC14C mutations.
C) KM survival analysis of PCAWG skin cancer patients stratified by the presence or absence of rare non-silent TUBA3D mutations.
D) KM survival analysis of PCAWG ovarian cancer patients stratified by the presence or absence of rare non-silent PI4KA mutations.

In all panels, mutation status = 0 indicates the presence and mutation status = 1+ indicates the absence of the rare non-silent mutations on the given gene.

### Literature Review on gene functions in cancer

| Gene | Cancer-Related Function Summary |
|---|---|
| **IRX3** | Oncogene in T-cell acute lymphoblastic leukemia (T-ALL); promotes tumorigenesis; upregulated in ~30% AML, ~50% T-ALL, 20% B-ALL; inhibits myeloid and T-cell differentiation[1-3]. |
| **ZNF717** | Limited data on cancer relevance. |
| **AOC1** | Promotes tumor progression via ferroptosis modulation and AKT/EMT signaling; upregulated in prostate, gastric, and colorectal cancers[4-6]. |
| **MUC12** | Independent prognostic marker in colorectal cancer; oncogenic in renal cell carcinoma; immunosuppressive and prognostic in hepatocellular carcinoma[7-10]. |
| **HRNR** | Promotes tumor progression and poor prognosis in hepatocellular carcinoma; potential therapeutic target; prognostic marker in gastric and colorectal cancer[11-13]. |
| **OR6B2** | Limited data on cancer relevance. |
| **CDC14C** | Involved in cell cycle regulation[14,15]. |
| **PRAMEF1** | PRAME family member, highly expressed in various cancers and is a preferentially expressed antigen of melanoma[16]. |
| **MST1L** | Identified as a metastasis-specific mutated gene in Chinese colorectal cancer[10]. |
| **FOLH1** | Prognostic marker in renal cell carcinoma; high expression correlates with improved overall survival in metastatic tumors[17,18]. |
| **KRTAP9-2** | Copy number variation observed in urothelial carcinoma clinical trial[19]. |
| **TUBA3D** | Frequently mutated in BRCA; associated with platinum resistance and taxane resistance in basal-like tumors[20-22]. |
| **PI4KA** | Sensitizes refractory leukemia to chemotherapy; prognostic marker in prostate cancer; involved in bone tumor growth[23-25]. |
| **HLA-DRB1** | Key immune gene; marker for cancer risk and prognosis[26-28]. |
| **EPPK1** | Prognostic biomarker in endometrial cancer; promotes proliferation in cervical cancer; linked to EMT, cancer progression, and smoking status[29-31]. |

1. Rahman, S. *et al.* IRX3 Oncogene Activation in T-ALL through Enhancer

Hijacking Caused By fto Intron 8 Deletions. (American Society of Hematology Washington, DC, 2022).
2. Wang, P., Zhuang, C., Huang, D. & Xu, K. Downregulation of miR-377 contributes to IRX3 deregulation in hepatocellular carcinoma. *Oncology reports* **36**, 247-252 (2016).
3. Somerville, T.D. *et al.* Derepression of the Iroquois homeodomain transcription factor gene IRX3 confers differentiation block in acute leukemia. *Cell Reports* **22**, 638-652 (2018).
4. Ding, Y. *et al.* SOX15 transcriptionally increases the function of AOC1 to modulate ferroptosis and progression in prostate cancer. *Cell death & disease* **13**, 673 (2022).
5. Xu, F. *et al.* AOC1 contributes to tumor progression by promoting the AKT and EMT pathways in gastric cancer. *Cancer Management and Research*, 1789-1798 (2020).
6. Liu, F. *et al.* Increased AOC1 expression promotes cancer progression in colorectal cancer. *Frontiers in Oncology* **11**, 657210 (2021).
7. Matsuyama, T. *et al.* MUC12 mRNA expression is an independent marker of prognosis in stage II and stage III colorectal cancer. *International journal of cancer* **127**, 2292-2299 (2010).
8. Gao, S.L. *et al.* The oncogenic role of MUC12 in RCC progression depends on c-Jun/TGF-β signalling. *Journal of Cellular and Molecular Medicine* **24**, 8789-8802 (2020).
9. Huang, H., Hu, Y., Guo, L. & Wen, Z. Integrated bioinformatics analyses of key genes involved in hepatocellular carcinoma immunosuppression. *Oncology Letters* **22**, 830 (2021).
10. Yang, Y. *et al.* Mutational profile evaluates metastatic capacity of Chinese colorectal cancer patients, revealed by whole-exome sequencing. *Genomics* **116**, 110809 (2024).
11. Fu, S.-J. *et al.* Hornerin promotes tumor progression and is associated with poor prognosis in hepatocellular carcinoma. *BMC cancer* **18**, 815 (2018).
12. Dimastromatteo, J. & Kelly, K.A. Hornerin as a novel therapeutic target for pancreatic cancer. *Cancer Research* **79**, 3049-3049 (2019).
13. Oshima, T. *et al.* Clinical significance of HRNR expression in patients with stage II/III gastric cancer after curative gastrectomy. *Anticancer Research* **44**, 4579-4584 (2024).
14. Manzano-López, J. & Monje-Casas, F. The multiple roles of the Cdc14 phosphatase in cell cycle control. *International Journal of Molecular Sciences* **21**, 709 (2020).
15. Roy, S.H., Clayton, J.E., Holmen, J., Beltz, E. & Saito, R.M. Control of Cdc14 activity coordinates cell cycle and development in Caenorhabditis elegans. *Mechanisms of Development* **128**, 317-326 (2011).
16. Nettersheim, D. *et al.* The cancer/testis-antigen PRAME supports the pluripotency network and represses somatic and germ cell differentiation programs in seminomas. *British journal of cancer* **115**, 454-464 (2016).
17. Royz, E. *et al.* Characterization of FOLH1 expression in renal cell carcinoma. *Cancers* **16**, 1855 (2024).
